## AppendixS1 for "Short-term outcomes of pubertal suppression in a selected cohort of 12 to 15 year old young people with persistent gender dysphoria in the UK"

APPENDIX S1

S1 Search Terms

Search terms for review conducted in medline (www.pubmed.com) on 2-1-2020

((((((((GD) OR transsexualism) OR ("sexual and gender disorders")) OR transgender persons) OR gender identity) OR gender dysphoria)) AND (((((("drug therapy"[MeSH Terms]) OR "therapeutic uses"[MeSH Terms]) OR hormone*) OR "steroids"[MeSH Major Topic]) OR gestagen) OR antiandrogens[MeSH Terms])) AND (((adolescent[MeSH Terms]) OR teen*) OR "paediatric")

S1 Table S1. Summary of Interview data at 6-15 months

The table shows proportions rating change in each of the domains as being either positive, negative, both positive and negative, no change or not known.

For sexuality, change was not accorded a positive or negative valency, and is reported as ‘any change’ compared with ‘no change’.

| n=41 | Life overall | Memory | Focus | Direction | Mood | Energy | Family | Friends | Gender role | Sexuality | Other experiences |
|---|---|---|---|---|---|---|---|---|---|---|---|
| Positive | 19 (46%) | 2 (5%) | 2 (5%) | 0 (0%) | 20 (49%) | 2 (5%) | 10 (24%) | 10 (24%) | 27 (66%) | - | 5 (12%) |
| Both positive & negative | 15 (37%) | 0 (0%) | 0 (0%) | 0 (0%) | 6 (15%) | 3 (7%) | 5 (12%) | 3 (7%) | 0 (0%) | Any change 2 (5%) | 4 (10%) |
| No change | 2 (5%) | 33 (80%) | 33 (80%) | 39 (95%) | 3 (7%) | 23 (56%) | 19 (46%) | 25 (61%) | 13 (32%) | 35 (85%) | 10 (24%) |
| Negative | 5 (12%) | 5 (12%) | 6 (15%) | 0 (0%) | 10 (24%) | 12 (29%) | 3 (7%) | 3 (7%) | 0 (0%) | - | 0 (0%) |
| Not known | 0 (0%) | 1 (2%) | 0 (0%) | 2 (5%) | 2 (5%) | 1 (2%) | 3 (7%) | 0 (0%) | 0 (0%) | 4 (10%) | 0 (0%) |
| Not completed | 0 (0%) | 0 (0%) | 0 (0%) | 0 (0%) | 0 (0%) | 0 (0%) | 1 (2%) | 0 (0%) | 1 (2%) | 0 (0%) | 22 (54%) |

S1 Table S2. Summary of Interview data at 15-24 months

The table shows proportions rating change in each of the domains as being either positive, negative, both positive and negative, no change or not known.

For sexuality, change was not accorded a positive or negative valency, and is reported as 'any change' compared with 'no change'.

| n=29 | Life overall | Memory | Focus | Direction | Mood | Energy | Family | Friends | Gender role | Sexuality | Other experiences |
|---|---|---|---|---|---|---|---|---|---|---|---|
| Positive | 16 (55%) | 1 (3%) | 3 (10%) | 1 (3%) | 8 (28%) | 2 (7%) | 13 (45%) | 10 (34%) | 12 (41%) | - | 10 (34%) |
| Both positive & negative | 5 (17%) | 0 (0%) | 0 (0%) | 0 (0%) | 8 (28%) | 0 (0%) | 1 (3%) | 3 (10%) | 0 (0%) | Any change 6 (21%) | 1 (3%) |
| No change | 3 (10%) | 26 (90%) | 19 (66%) | 26 (90%) | 3 (10%) | 15 (52%) | 13 (45%) | 14 (48%) | 17 (59%) | 21 (72%) | 11 (38%) |
| Negative | 5 (17%) | 1 (3%) | 6 (21%) | 1 (3%) | 8 (28%) | 11 (38%) | 1 (3%) | 1 (3%) | 0 (0%) | - | 6 (21%) |
| Not known | 0 (0%) | 0 (0%) | 1 (3%) | 1 (3%) | 1 (3%) | 1 (3%) | 1 (3%) | 1 (3%) | 0 (0%) | (0%) | 0 (0%) |
| Not completed | 0 (0%) | 1 (3%) | 1 (3%) | 0 (0%) | 1 (3%) | 0 (0%) | 0 (0%) | 0 (0%) | 0 (0%) | 2 (7%) | 1 (3%) |

S1 Appendix Table S3. Sensitivity analyses of ASEBA outcome testing restricted to rescored data

| | | | **12 months** | Baseline in those with data at 12m | Change from baseline to 12 months | p | | **24 months** | Baseline in those with data at 24m | Change from baseline to 24 months | p |
|---|---|---|---|---|---|---|---|---|---|---|---|
| | | n | mean (95% CI) | mean (95% CI) | mean (95% CI) | | n | mean (95% CI) | mean (95% CI) | mean (95% CI) | |
| Parent report CBCL | Total problems t-score | 40 | 62.0(58.5, 65.4) | 61.5(58.1, 64.9) | 0.5(-1.9, 2.8) | 0.7 | 20 | 60.2(54.6, 65.8) | 61.2(56.5, 65.8) | -1.0(-4.0, 2.1) | 0.5 |
| Self-report YSR | Total problems t-score | 38 | 58.6(54.5, 62.7) | 57.4(54.2, 60.6) | 1.2(-3.0, 5.4) | 0.5 | 14 | 56.7(50.7, 59.7) | 55.2(50.7, 59.7) | 1.5(-3.7, 6.7) | 0.6 |
| **Self-harm scores** | | | | | | | | | | | |
| Parent report CBCL | Median (IQR) | 40 | 0(0, 1) | 0(0, 1) | 0.3 | | 20 | 0(0, 1) | 0(0, 1) | | 1.0 |
| Self-report YSR | Median (IQR) | 37 | 0(0, 2) | 0(0, 1) | 0.4 | | 15 | 0(0, 0) | 0(0, 1) | | 1.0 |

| **36 month data** | | | **36 months** | **Baseline in those with data at 36m** | **Change from baseline to 36 months** | **p** |
|---|---|---|---|---|---|---|
| | | n | mean (95% CI) | mean (95% CI) | mean (95% CI) | |
| Parent report CBCL | Total problems t-score | 11 | 61.1(52.3, 69.9) | 62.4(55.1, 69.6) | -1.3(-6.6, 4.0) | 0.6 |
| **Self-harm scores** | | | | | | |
| Parent report CBCL | Median (IQR) | 11 | 0(0, 1) | 0(0, 1) | | 0.8 |

### S1 Semi-structured interview questions

Question 1- Have you noticed any changes in your life since starting on the hormone blocker

Question 2- Have you noticed any changes in how you remember things since starting on the hormone blocker?

Question 3- Have you noticed any differences in how you are able to focus on things since starting on the hormone blocker?

Question 4- Have you noticed any difference in finding your way to go somewhere or directing other people to go somewhere since starting on the hormone blocker?

Question 5- Has your mood or how you are feeling changed since starting on the hormone blocker?

Question 6- 6. Have your energy levels changed since starting on the hormone blocker?

Question 7- Have you had any other experiences that we haven't mentioned since starting on the hormone blocker?

Question 8- Has your relationship with your family members changed since starting on the hormone blocker?

Question 9- Has your gender role (in terms of living more as a male or a female) changed since starting on the hormone blocker?

Question 10- Has your relationship with your friends or peers changed since starting on the hormone blocker?

Question 11- How would you describe your sexuality at the moment? (E.g. are you attracted to females, males, both females and males, not attracted to either or not sure? Is that different from before you started on the blocker?

Question 12- Do you want to continue on the blocker.
