## Appendix S2 for "Short-term outcomes of pubertal suppression in a selected cohort of 12 to 15 year old young people with persistent gender dysphoria in the UK"

### S2 Appendix: Statistical Analysis Plan.

Version 1.0, 9 October 2019

The following document was lodged with the Research Ethics Committee of the Health Research Authority on 9 October 2019, before analysis commenced in this study.

#### Baseline assessment

Eligibility criteria were assessed at recruitment. They will be reported in summary form i.e. pubertal stage and phenotype of genitalia, bone age, and normality of karyotype, pelvic ultrasound and short synacthen test (the latter two for birth-registered females only).

#### Outcomes

Outcomes at baseline and follow-up were assessed in four domains:

a. Puberty and growth
b. Medical safety
c. Bone health
d. Psychological function

Two outcomes were assessed only during or after treatment.

e. Treatment satisfaction
f. Decision made at the end of treatment.

The Table below lists the specific outcomes from each domain, along with their expected or hypothesised change from baseline and the analytic plan for each outcome.

| Outcome | Description and measurement | Timing | Expected or hypothesised change on GnRHa | How assessed for study outcomes |
|---|---|---|---|---|
| A. PUBERTY & GROWTH | | | | |
| 1. Gonadotropins and sex steroids | Gonadotropins (LH, FSH; in both sexes), testosterone (in birth-registered males) and oestrogen (birth-registered females) | At 6 and 12 months then annually | The aim of GnRHa treatment was to suppress LH and FSH production, leading to the 'turning off' of gonads and therefore suppression of testosterone or oestrogen (depending on birth registered sex).<br><br>Hypothesis: that hormones will be suppressed to prepubertal levels by 6 to 12 months. | a. Categorical: Report of whether sex steroids and gonadotropins were suppressed at 6 to 12 months<br>b. Mean value and 95% CI for hormones at 12, 24 and 36 months |
| 2. Growth | Height, weight and calculated BMI | 6 monthly initially and thereafter 12-monthly | Height: Height growth during puberty is dependent on the twin effects of sex steroids (oestrogen or testosterone) and growth hormone. The pubertal growth spurt occurs around Tanner stage 2-3 in females and stage 3-4 in males. GnRHa treatment is expected to dramatically reduce height growth compared to adolescent peers who are growing rapidly. This will show as only small growth in height but a fall in height z-score (i.e. standard deviation score adjusted for age and sex). | z-scores for height, weight and BMI calculated for age and sex at 12, 24 and 36 months using the UK 1990 growth reference. Analysis will be restricted to calculation of means and 95% confidence intervals (95% CI) at each time point, and no statistical testing will be done |

| Outcome | Description and measurement | Timing | Expected or hypothesised change on GnRHa | How assessed for study outcomes |
|---|---|---|---|---|
| | | | Hypothesis: height velocity will be reduced to prepubertal levels, with a matching fall in height z-score.<br><br>Weight and BMI: No definitive hypotheses. We will examine weight and BMI standardized for age and sex through use of weight and BMI z-score. | |
| 3. Pubertal progress | Pubertal staging (Tanner staging) is conducted by physical examination or by self-report, and reports the degree to which young people have progressed from prepubertal (stage 1) to fully puberty (stage 5) in 3 areas for each birth registered sex.<br>a. genital and pubic hair stage and testicular volume in males;<br>b. Breast and pubic hair stage and menarcheal status in females. | Baseline and end of follow-up | The aim of GnRHa treatment is to prevent further progression in puberty during treatment. Young people will be categorised as stage 2 through 5 at baseline and monitored for pubertal progression.<br><br>Hypothesis: No pubertal progression | Baseline: proportions in each Tanner stage<br><br>Follow-up: proportions with any pubertal progression or not |

| **Outcome** | **Description and measurement** | **Timing** | **Expected or hypothesised change on GnRHa** | **How assessed for study outcomes** |
|---|---|---|---|---|
| | For analysis the three values for each birth registered sex were combined into an overall stage i.e. 2 through 5. This is common practice in clinical research. | | | |
| B. MEDICAL SAFETY | | | | |
| 1. Safety bloods | 1. Liver & renal function<br>2. Full blood count<br>3. Bone profile<br>4. Basal endocrinology<br>5. Vitamin D | 12-monthly.<br><br>Liver and renal function also assessed at 3m. | Safety bloods were designed to assess impact on liver and renal function, basal endocrinology and basic haematology.<br><br>Hypotheses: No change in any measured parameters | Individual tests are not outcomes of interest aside from overall safety. Bloods were taken at regular intervals but for parsimony will reported at 12, 24 and 36 months. Reporting of outcomes will be categorical in terms of normality of each test, with numbers with identified abnormalities of any test. The reference norms for the laboratory where testing was done (UCLH) will be used to |

| Outcome | Description and measurement | Timing | Expected or hypothesised change on GnRHa | How assessed for study outcomes |
|---|---|---|---|---|
| | | | | define normality. No statistical testing will be done. |
| 2. Adverse events | Patient report of adverse events from<br>a. clinic visits<br>b. patient interviews<br>At each clinic visit, patients were routinely asked about adverse events. | At 6, 12, 24, 36 and 48m | Expected adverse effects were not specified in the Protocol. Any adverse events noted at clinic visits will be reported.<br><br>Note that formal semi-structured interviews at 6-12 months after commencement also sought information on side-effects and these data will be contribute to side-effect reporting. | Presence or absence of reported side-effects |
| C. BONE HEALTH | | | | |
| 1. Bone mineral density | DEXA scan report of bone mineral density in lumbar spine (L1-4) and hip:<br>Primary outcomes in spine and hip:<br>1. Bone mineral content (BMC)<br>2. Bone mineral density (BMD)<br>Secondary outcomes: calculated BMD z-score for age & sex, regression-adjusted for change in height z-score (height adjusted z-score: HAZ) | Annually | There is a strong relationship between bone strength, height and puberty. Normatively, BMC and BMD rise during adolescence due to the action of sex steroids on bone mineral accretion and because of height growth.<br><br>This will not occur if puberty is suppressed, as height does not increase and because bone mineral accretion | Change in continuous measures of BMC, BMD and BMD HAZ at 12, 24 and 36 months.<br><br>Formal statistical testing will be restricted to change in BMC and BMD as primary outcomes. Means and 95% confidence |

| Outcome | Description and measurement | Timing | Expected or hypothesised change on GnRHa | How assessed for study outcomes |
|---|---|---|---|---|
| | | | is not promoted by sex steroid. The expectation is therefore that during GnRHa treatment bone strength as measured by BMC and BMD will not fall during treatment – but that it will not increase as expected from age and height growth. BMD z-scores for age reflect population increases in bone mineral, therefore z-scores will fall if puberty is suppressed, although without any reduction in bone strength.<br><br>We hypothesise that there will be no change in absolute BMD nor in BMC. However we expect that BMD z-score will fall consistent with falls in height z-score i.e. that HAZ will remain constant. | intervals (95% CI) will be calculated for HAZ at each time point. |
| D. PSYCHOLOGICAL FUNCTION | | | | |
| 1. Child Behavior Checklist (CBCL) | The Child Behavior Checklist (CBCL) (parent report) and Youth Self Report (YSR) (self-report) are measures of psychological functioning | At baseline and again | Normative data show an increase in CBCL internalizing scores (i.e. psychological distress) with age in early adolescence.[22] It is therefore likely that any | We will use t- scores for comparison with normative published data: |

| Outcome | Description and measurement | Timing | Expected or hypothesised change on GnRHa | How assessed for study outcomes |
|---|---|---|---|---|
| and Youth Self-Report (YSR) | (aseba.org). Both provide continuous data/ t-scores for the total problem scale (a global index of functioning across all subscales). They also provide continuous data/ t-scores on the internalizing subscale (items which assess anxious/depressed, withdrawn-depressed, and somatic complaints) and the externalizing subscale (focusing on rule-breaking and aggressive behaviours). The CBCL is used for children aged 6 to 18 and consists of 113 questions. Both are scored on a three-point Likert scale (0=absent, 1= occurs sometimes, 2=occurs often). The time frame for item responses is the past six months.<br><br>Raw scores are converted for analysis to t-scores based upon broad age-ranges.<br><br>Higher scores indicate greater morbidity. | after around 12 months on treatment. | improvements in psychological function with treatment are likely to result in smaller rises in distress with age, meaning that there will be no discernible change in psychological function over time in our cohort.<br><br>Previous data from the London service and internationally show high rates of psychological distress in young people with GID.[42] Thus our sample is a group in whom distress begins at levels higher than the general population, but who will also suffer from the normative increase in distress across the study period.<br><br>We therefore hypothesise there will be<br>1. no change in total problem t-scores<br>2. no change in externalizing or internalizing subscale t-scores | 1. total problems score<br>2. internalising subscale score<br>3. externalising subscale score<br>Change in total problems score will be tested. Change in internalising and externalising t-scores will be estimated by calculating mean and 95% CI. |

| Outcome | Description and measurement | Timing | Expected or hypothesised change on GnRHa | How assessed for study outcomes |
|---|---|---|---|---|
| | | | This is consistent with a potential reduction in distress due to treatment at a time when there is a developmental rise in distress. | |
| 2. Youth Self-Report (YSR) | The YSR is the self-report version of the CBCL (see above). The YSR is suitable for children and adolescents aged 11-18 years and consists of 112 questions.<br><br>Higher scores indicate greater morbidity.<br><br>As per the CBCL, raw scores are converted to t-scores for analysis. However t-scores are based upon broad age-ranges. To account for normative change across our age-range, we will also use international reference data[23] to transform raw scores into z-scores for age and sex. | Baseline and follow-up | Normative data show rising YSR total problems scores with age from age 11 to 16 years in healthy young people from a range of different countries.[23]<br><br>As above we hypothesize that there will be<br>1. no change in total problem t-scores<br>2. no change in z-scores for total problems<br>3. no change in externalizing or internalizing subscale scores | Change in total problems t-score will be tested. Change in internalising and externalising t-scores will be estimated by calculating mean and 95% CI at each time point. Change in total problems z-scores will be estimated by calculating mean and 95% CI at each time point. |
| 3. Self-harm | This will be assessed by 2 questions in each of the CBCL (parent report) and YSR (self-report). | | Self-harm rates in the general population in the UK and elsewhere increase markedly with age across early to mid-adolescence, from very low in <10 year | We will assess change by change in self-harm/suicide items score. This score combines all categories |

| **Outcome** | **Description and measurement** | **Timing** | **Expected or hypothesised change on GnRHa** | **How assessed for study outcomes** |
|---|---|---|---|---|
| | Item 18: I deliberately try to hurt or kill myself (self-harm)<br>Item 91. I think about killing myself (suicidal ideation)<br><br>For each question in both CBCL and YSR the responses are:<br>0=not true<br>1=somewhat or sometimes true<br>2= very true or often true<br><br>We will use these items to calculate a self-harm/suicide items score at each timepoint by sum of the 2 items in each of the CBCL and YSR, as published by other authors.[24, 25] This produces a scale from 0 to 4 for each question – with higher score indicating greater suicidality/self-harm thoughts and behaviour. | | olds to peak around age 16-17 years.[43-46] This is likely related to adolescent development, particularly cognitive development.<br><br>Self-harm is highly prevalent in young people with GID, similar to rates in young people with a range of serious mental health problems.[34]<br><br>Thus our sample is a group in whom self-harm begins at levels higher than the general population, but who will also suffer from the normative increase in self-harm across the study period.<br><br>We hypothesise no change in self-harm across the study. This is consistent with a reduction in the propensity to self-harm due to treatment at a time when there is a developmental rise in self-harm. | of self-harm and suicidal ideation, including intermediate and high categories of risk. This will be formally tested. |

| Outcome | Description and measurement | Timing | Expected or hypothesised change on GnRHa | How assessed for study outcomes |
|---|---|---|---|---|
| 4. Teacher Report Form | The Teacher Report Form (TRF) is the third element of the Achenbach ASEBA system, asking teachers similar questions to the CBCL (parents) and YSR (young person self-report). Attempts to obtain TRF data on participants were rarely successful as this relied on families organising completion and return of questionnaires. Sample size for the TRF are insufficient to report, therefore data are not available on this outcome. | | | Not assessed. |
| 5. Children's Global Assessment Scale | The Children's Global Assessment Scale (CGAS), adapted from the Global Assessment Scale for adults, is a rating of functioning aimed at children and young people aged 6-17 years old. The child or young person is given a single score between 1 and 100, based on a clinician's assessment of a range of aspects related to a child's psychological and social functioning, with the time period being the last month. The score puts them in one of ten categories that range | | Hypothesis: improvement in CGAS score with treatment. | Change will be estimated by calculating mean and 95% CI. |

| Outcome | Description and measurement | Timing | Expected or hypothesised change on GnRHa | How assessed for study outcomes |
|---|---|---|---|---|
| | from ‘extremely impaired’ (1-10) to ‘doing very well’ (91-100).[29] The CGAS is a commonly used measure in child and adolescent mental health services in the UK.<br><br>Higher scores indicate better function. | | | |
| 6. Kidscreen- 52 | Separate young person and parent questionnaires each consist of 52 items which assess ‘Health Related Quality of Life’ (HRQoL) across ten dimensions: physical well-being; psychological well-being; moods and emotions; self-perception; autonomy; relations with parents and home life; social support and peers; school environment; social acceptance (bullying); and financial resources. Items use five-point Likert-style scales to assess either the frequency (never-seldom-sometimes-often-always) of certain behaviours/feelings or the intensity of an attitude (not at all–slightly- | | As for other items, It is likely that many of the Kidscreen subscale scores will reflect normative reductions in wellbeing and rises in mental health problems across early adolescence.<br><br>Hypothesis: no change in sub-scales. | Change in parent and child subscales will be estimated by calculating means and 95% CI. |

| Outcome | Description and measurement | Timing | Expected or hypothesised change on GnRHa | How assessed for study outcomes |
|---|---|---|---|---|
| | moderately-very-extremely). The measure was developed for young people aged 8-18 years. The recall period is one week.<br><br>The questionnaires provide scores in the form of continuous t-scores for the 10 subscales. T-scores are derived from a multinational European sample.[27]<br><br>Lower scores indicate greater morbidity. | | | |
| 7. Body Image Scale (BIS) | The BIS is a self-report measure used to assess body image satisfaction/ dissatisfaction. The questionnaire provides a total score in the form of continuous data/ a t-score for the main scale (sum of all subscales) as well as for three subscales assessing 'primary', 'secondary' and 'neutral' characteristics separately.[28]<br><br>The questionnaire is used for children and adolescents aged 12 years and over. The | | Young people with GID have significant dissatisfaction with their bodies, particularly those sexually dimorphic elements.[47]<br><br>GnRHa treatment does not change the body in terms of becoming the desired gender. GnRHa treatment only stops further progression in puberty i.e. it stops further development of unwanted gender characteristics. It is therefore unlikely that GnRHa treatment will result in significant reduction in body | Change in t-scores for main/total scale and for the subscales (primary, neutral and secondary sex characteristics) will be estimated by calculating mean and 95% CI. No formal statistical testing will be undertaken. |

| Outcome | Description and measurement | Timing | Expected or hypothesised change on GnRHa | How assessed for study outcomes |
|---|---|---|---|---|
| | measure consists of 30 body features which the respondent is asked to rate in terms of satisfaction on a five-point scale (1= very satisfied, 2 = satisfied, 3= neutral, 4 = dissatisfied, and 5 = very dissatisfied). The measure provides three scores for 'primary', 'secondary' and 'neutral' characteristics. The primary characteristics subscale describes satisfaction/dissatisfaction with the penis, scrotum, testicles, facial hair, body hair, and breasts for assigned males, and vagina, clitoris, ovaries-uterus, breasts, chest, facial hair, and voice for assigned females. The secondary characteristics subscale is comprised of hips, figure, waist, arms, buttocks, biceps, appearance, stature, muscles, weight, thighs, and hair for both sexes. It also includes voice and chest for assigned males and body hair for assigned females. The 'neutral' subscale, defined as 'neutral' as the authors considered | | dissatisfaction.[13] Use of cross-sex hormones appears to improve body dissatisfaction in those with GID[47].<br><br>Hypothesis: There will be no change in BIS scores with GnRHa treatment. | |

| Outcome | Description and measurement | Timing | Expected or hypothesised change on GnRHa | How assessed for study outcomes |
|---|---|---|---|---|
| | them unresponsive to hormonal interventions, is comprised of nose, shoulders, chin, calves, hands, Adam's apple, eyebrows, face, feet, and height for both sexes.<br><br>Higher scores represent higher degrees of body dissatisfaction. | | | |
| 8. The Utrecht Gender Dysphoria Scale (UGDS) | The GDS is a self-report measure used to assess the intensity of Gender Dysphoria. The questionnaire provides a total score in the form of continuous data/ a t-score for one main scale only.<br><br>The questionnaire is used for children and adolescents aged 12 years and over. The questionnaire consists of 12 statements and answers are given on a five-point scale to indicate to what extent one agrees or disagrees (1= agree completely, 2 = agree somewhat, 3= | | As noted above, GnRHa treatment does not change the body but merely prevents further pubertal changes. GnRHa treatment is therefore unlikely to affect feelings of gender dysphoria.<br><br>Hypothesis: gender dysphoria does not change with GnRHa treatment | Change in t-scores for total scale will be estimated by calculating mean and 95% CI. No formal statistical testing will be undertaken. |

| **Outcome** | **Description and measurement** | **Timing** | **Expected or hypothesised change on GnRHa** | **How assessed for study outcomes** |
|---|---|---|---|---|
| | neutral, 4 = disagree somewhat, and 5 = disagree completely). A maximum score of 60 can be obtained. There are two separate versions of the UGDS for assigned males (UGDS-M) and assigned females (UGDS-F)<br><br>Higher scores indicate greater morbidity. | | | |
| 9. Social Responsiveness Scale | The Social Responsiveness Scale (SRS) (school form) is a quantitative measure of autistic traits and symptoms in 4–18 year olds. It includes 18 items. A total score is produced.[59] | | Young people with GID have been described to have relatively high levels of autistic traits. This may affect the natural history of and response to treatments for GID. It is unclear whether autistic traits would influence response to GnRHa treatment.<br><br>The SRS is used here as a baseline assessment of autistic traits and as a predictor of outcome and NOT as an outcome measure itself. The SRS was assessed only at baseline and not at follow-up as it was not anticipated that scores would change. | Not applicable. |

| Outcome | Description and measurement | Timing | Expected or hypothesised change on GnRHa | How assessed for study outcomes |
|---|---|---|---|---|
| E. Satisfaction with treatment | | | | |
| 1. Satisfaction with treatment<br><br>2. Continuation of treatment | Satisfaction with treatment was assessed in semi-structured interviews 6-12 months after commencing treatment.<br><br>Whether young people and families wished to continue on treatment was assessed at each clinic visit | 6 and 12 months on treatment<br><br>Each clinic visit | No hypothesis appropriate. No baseline data available. | Descriptive proportions.<br><br>Descriptive proportions |
| F. Decisions made at end of GnRHa treatment pathway | | | | |
| | GnRHa treatment was only provided in the absence of sex steroids until around 16 years. After age 16 years young people were eligible to elect to start cross-sex hormones. Young people made a decision at the end of the | After age 16 years. | No hypothesis. | Descriptive proportions. |

| Outcome | Description and measurement | Timing | Expected or hypothesised change on GnRHa | How assessed for study outcomes |
|---|---|---|---|---|
| | GnRHa pathway to either cease GnRH (with the result that their body resumes production of sex steroids) or commence cross-sex hormones. This is a binary decision, made after at least 12 months on GnRHa and after the young person's 16th year. | | | |

#### Analysis methods

The original protocol stated: "All the data will be recorded and the questionnaires scored. Descriptive statistics including means and standard deviations will be reported as well as parametric and non-parametric tests as appropriate."

Due to the small sample size, analyses will be largely restricted to descriptive statistics and elementary statistical tests.

Outcomes will be reported at baseline and 12, 24 and 36 months. Outcomes at 48 months will not be reported due to very low numbers.

For categorical outcomes, simple descriptive statistics will be reported.

For continuous measures we will formally test change using paired t-tests for the significance of the before/after difference of the main scales as specified above. For other scales and subscales we will calculate means and 95% CI at baseline and follow-up, showing baseline mean for both the whole sample and for the sample with data at the follow-up point.

Predictors of outcome: There are multiple hypotheses that could be made about whether individual demographic, physical or psychological factors relate to differences in outcomes, either by confounding or potentially more causally. Our design and study size mean that exploration of such hypotheses is not possible without high risk of chance findings. We will restrict analysis of predictors of outcome to two factors, birth registered sex and pubertal stage at baseline. We will restrict analysis to three continuous outcomes measured at baseline and 12 months. These outcomes were chosen as key outcomes across physical and mental health:

a. bone mineral density (BMD) in the lumbar spine
b. YSR total t-score
c. CGAS score

We will do this by linear regression of follow-up score on baseline score, adding each predictor separately to each model and considering the interaction of predictor with baseline score. If both factors appear to predict outcome, we will include these in a multivariable model and use backwards stepwise selection to identify the best predictive model.

We note that the nature of the patient group and their data are both unusual and highly sensitive, so that publication of findings may potentially identify individuals, particularly given the small sample size. We will consult with data protection experts to ensure that personal data are not disclosed.

#### Dissemination and Outputs

The findings of this study are of considerable public interest. They will first be published in a peer-reviewed international journal with the paper made open-access. They will be also presented at academic and practitioner meetings. In addition, the investigators will seek to share the findings more widely with patient groups and other stakeholders, using plain English.

The Tavistock will prepare a patient information sheet summarising findings for participants and families and circulate it to the participants and their GPs.
